# Using traditional rhyme (folk song) as a tool for oral hygiene promotion (UTRATOHP) among children in rural communities in Nigeria: a protocol for a randomised controlled trial

**DOI:** 10.1101/2023.01.11.23284418

**Authors:** Omotayo F. Fagbule, Urenna Emenyonu, Ejiro Idiga, Olubunmi O. Oni, Olabode A. Ijarogbe, Mary E. Osuh, Folake B. Lawal, Tolulope O. Owoaje, Olushola Ibiyemi

## Abstract

**Background:** Most oral diseases are caused by poor oral hygiene, and with adequate oral hygiene measures, they are easily preventable. The use of folk songs to deliver oral health education may likely hold a huge potential for success and an increased likelihood of acceptability and sustainability among school children. Therefore, an investigation into the effectiveness of methods that would be culturally appropriate and sustainable, such as folk songs, is essential.

**Aim:** To determine the effect of traditional rhyme (folk song) as a tool for oral hygiene education among children in rural communities in Nigeria.

**Materials and methods:** This is a school-based, assessor-blinded, two-arm cluster-randomised controlled trial that will assess the effectiveness of oral hygiene messages delivered through traditional rhyme (folk song) on children’s oral hygiene knowledge, attitude, practices, and oral hygiene status in two rural communities (Igboora and Idere) in Nigeria. The study will involve a minimum of 424 participants (aged 7-9 years) selected from eight primary schools using the cluster sampling technique. Four schools will be randomised into the test group to receive oral hygiene messages through folk songs, and the participants will sing the song for two weeks. The other four schools will be assigned to the control group, and the participants will receive the usual oral health talk on oral hygiene practices by a dentist.

The participants’ oral hygiene knowledge, attitude, practice and status will be assessed at baseline, immediate, six, and twelve-month post-intervention. A modified World Health Organization (WHO) Children’s oral health survey questionnaire will be utilised for data collection. Oral examinations will be conducted to assess the participants’ dental caries experience and oral hygiene status using the dmft/DMFT and simplified oral hygiene indices, respectively.

**Discussion:** Folk songs are popular means of conveying messages in Nigeria, and using them to deliver oral health messages may be an effective, acceptable, and sustainable method among children. This study will provide empirical information about this innovative intervention to guide policy development, dental public health practice, and future studies.

**Trial registration:** Pan African Clinical Trial Registry-PACTR202010863892797 (October 2020)

## Introduction

Oral diseases remain major public health problems with a substantial personal, social and economic burden.[1] Worldwide, the prevalence of oral diseases like dental caries and periodontal diseases is high as they affect most people in high-middle- and low-income countries (HICs and LMICs).[1] It is estimated that 2.4 billion people suffer from dental caries of permanent teeth, and 486 million children suffer from dental caries of primary teeth.[2] The global increase in the prevalence of dental caries[3,4] highlights the need for primary prevention programmes, especially in LMICs, including sub-Saharan Africa, which has the highest burden of untreated dental caries.[1,5] Dental caries are painful, negatively impact the quality of life, and may result in tooth loss and systemic infection that can lead to death in its advanced stage.[6] The treatment options for dental caries are costly, averaging 5% of the total health expenditure and 20% of out-of-pocket expenditure in developed countries.[7] In most developing countries, investment in oral health is low, and if treatment were available, the costs of dental caries in children alone would exceed the total health care budget.[8] Similarly, periodontal diseases contribute significantly to the global burden of oral disease, leading to pain, bleeding, bad breath, tooth loss and adverse systemic illnesses.[9] Most epidemiological studies have observed significant relationships between socio-economic status and periodontal disease.[10–13] Low income and low education contribute to poor periodontal disease status.[10–12] There is an unequal burden of oral diseases affecting those in the rural areas more than those in the urban communities,[13] and more among those living in slums than non-slum areas.[12]

Most oral diseases like dental caries and periodontal problems are preventable.[9] Dental caries is one of the most common oral diseases that negatively impact the oral health-related quality of life of those affected.[14] And while it can affect people over their life course,[15] the risk is higher among children.[15] Given the cost of treatment and devastating effect on the quality of life, prevention of dental and other oral problems must be prioritised. The common preventive measures include maintaining good oral hygiene, eating a balanced diet with low refined-carbohydrate content, and regularly visiting the dental office for early detection and treatment of emerging oral problems.[16,17] It is advocated that oral health education programs are instituted among the populace, including those in the rural and underserved communities, to instil good hygiene practices among them.[13] It is even more helpful for these activities to be carried out among children who are just forming behaviours that affect their health.[18] Helping children develop the habit of brushing twice daily and maintaining good oral hygiene is key to a lifetime of healthy smiles. Thus, exploring the roles of innovative methods such as oral hygiene songs, particularly folk songs with an inherent cultural motif at developing and probably sustaining good oral hygiene habits among children becomes pertinent.

Oral hygiene songs are a great way to have children learn about and remember how to practice adequate oral health. Children and adolescents have fun while setting healthy lifetime habits when they learn through music. Tooth brushing songs set to familiar tunes make learning about oral health fun and easy. Melodies of traditional songs can be combined with words specifically adapted to make the song connect to a teachable concept. Parents, guardians and schoolteachers can use songs as a fun way to teach brushing and good oral hygiene. Using songs as a reminder that it is time for toothbrushing is something parents/guardians and their children/wards can do together daily. Although using folk songs to deliver oral health messages holds much promise, there is a lack of empirical information about its use among children in Nigeria. Hence, this study aims to determine the effect of traditional rhyme (folk song) as a tool for oral hygiene education among children in rural communities in Nigeria.

## Materials and methods

### Study design

The research is designed to be a cluster-randomised, assessor-blinded, controlled trial with two study groups – intervention and control. The randomised control design was adopted to provide a high level of evidence for the study.

### Study location and site

The planned study sites are Igboora and Idere, two rural communities in Oyo State, Southwestern Nigeria, about 7 kilometres apart. Farming and petty trading are common in the two communities, and Yoruba is the predominant tribe and language. The two communities have several primary schools. Most primary schools have classes from Primary one (Pry 1) to Primary six (Pry 6), while some end at Pry 5. Generally, most classes have five arms (A–E), and preliminary investigation shows that each class arm has an average of 25 pupils. The usual age of entry into primary school ranges between four to six years, and those in Pry 4 are usually between 7 – 9 years old.[19]

### Study population

The study participants would comprise Pry 4 pupils from primary schools located in the two rural communities (Igboora and Idere) in Oyo State, Nigeria. The choice of the Primary 4 class pupils is for the following reasons: the pupils’ ages range from seven to nine years, and they usually would have started supervised but unassisted oral hygiene practices like tooth brushing.[20,21] The age group (7-9 years) also has a higher likelihood of providing better responses during the interviews than younger people from the lower classes.[22] Moreover, pupils in the Pry 4 class are more likely to remain in their various schools through the proposed study period, unlike those in higher classes (Pry 5 and Pry 6) who may be nursing the idea of writing the secondary school qualifying examinations (common entrance examinations) and leaving their various schools while this study is still ongoing.

## Eligibility criteria

### Inclusion criteria

Healthy children with parental consent will be included in the study.

### Exclusion Criteria

Primary schools not registered with the Oyo State Government will be excluded from the study. Pupils who intend to leave the school before completing the study and those who do not understand the Yoruba language will also be excluded.

### Sample size determination

The sample size for this study was determined using the formula for comparing two proportions.[23] The primary outcome is oral hygiene status. The prevalence (P_1 =_ 38.4%) of children who had good oral hygiene from a previous study in Nigeria,[24] was used for sample size calculation. An estimated 50% increase in the proportion of participants with good oral hygiene following the intervention (P_2_ = 57.6%) was used in calculating the minimum sample size. Other considerations were statistical power of 80%, a significance level of 5%, a non-response rate of 10%, an attrition rate of 20%, and an estimated design effect of 1.5. A minimum of 212 participants will be recruited per study group, making 424 for the two groups.

### Sampling technique, recruitment, and randomisation

A two-stage cluster sampling technique will be employed for this study.

In the first stage, four schools from each of the two communities will be selected from the list of schools that would be obtained from the Ministry of Education, Oyo State, using the simple random sampling technique, making eight schools in total. In the second stage, an average of 55 participants will be selected from each selected school’s Pry 4 class arm using a proportionate sampling technique.

The schools will be the randomisation unit. The selected schools in the two communities (Igboora and Idere) will be randomised into either the test or the control group using the block randomisation method, aided by computer-generated numbers.[25] A community dentist, is unaware of the study details and the objectives, will allocate the schools and participants into the study groups.

### Baseline data collection

Interviewer-administered semi-structured questionnaire (S1 File) will be used for data collection. The questionnaire will be developed from the review of related local literature,[12,24,26] and the oral examination will be based on the WHO Oral Health Surveys: Basic methods, 5^th^ Edition.[27]. Five trained and calibrated dentists will conduct the interview and oral examinations. The dentists will be trained by an expert in oral epidemiology using the WHO guideline.[27] The questionnaire will be used to collect information on the participants’ sociodemographic characteristics, oral hygiene knowledge, attitude and practices. The oral examination will assess the participants’ oral hygiene status (simplified oral hygiene index[28]) and their lifetime caries experience [sum of decayed, missing, and filled teeth (dmft/DMFT)[27,29]]. The oral examination will be done using a sterile wooden mouth spatula, probes, and dental mirrors. The oral examination will be conducted near the windows in the school halls, with the participants seated upright facing the window such that the examination is aided by good illumination from natural light. Baseline data collection will be conducted in both study groups (test and control) one week before delivering the interventions.

### Intervention and control groups

The intervention will involve teaching the participants in the test group traditional rhyme (folk songs) on adequate oral hygiene measures and practices. The song was developed in English and Yoruba, the two major Nigerian languages in the two communities. While Yoruba is the primary language of the pupils, they are taught in English. The songs convey health messages, including the importance of twice-daily tooth brushing with pea-size fluoride-containing toothpaste, the need to brush the tongue, and simple tooth brushing techniques using toothbrushes with soft/medium-bristles. The song was co-developed and validated among selected schoolchildren, teachers, and parents/legal guardians.[30]

There will be three intervention sessions lasting for one month. During the first session, one of the researchers (OO) will teach selected participants in the test group the two songs (English and Yoruba). Some participants will be asked to sing the songs to confirm if the pupils have learned them. Subsequently, all the participants will be asked to sing the songs several times. The first session of the intervention is estimated to last for 40 minutes. The participants will be asked to continue singing the songs every morning in the school for one month, under the supervision of a teacher. This will be monitored by calling the designated teachers daily to confirm it was done. The second session will take place two weeks after the first, and the researcher will ask the participants to sing the songs as a group several times. Similarly, the third session will take place two weeks after the second session (one month after the first session) and the students will also sing the songs in the presence of the researcher (OO). The second and third sessions are estimated to last for 20 minutes.

The participants in the control group will be exposed to conventional health education on adequate oral hygiene practices conducted by the dentist.[31] Similar to the test group, there will be three intervention sessions. The first session will last for 40 minutes and will involve expert-led health talks and question and answer with the participants. The second and third sessions will also take place after two and four weeks, respectively. The follow-up sessions will involve revising the oral health messages with the participants, lasting for about 20 minutes each. None of the participants (control and test groups) will be denied access to oral health talks that they may be exposed to away from school.

### Post-intervention data collection

There will be three sessions of post-intervention assessments – Immediate, six and 12 months. The immediate post-intervention assessment will collect information on the participants’ oral hygiene knowledge and attitude immediately after the second session of the intervention is completed. The 6-month post-intervention assessment will be conducted six months after the immediate post-intervention assessment. Information on the participants’ oral hygiene knowledge, attitude, and practices will be collected. The 12-month post-intervention assessment will be conducted 12 months after the immediate post-intervention assessment. It will involve oral health examinations to check the participants’ oral hygiene status and dental caries experience (dmft/DMFT), along with assessing the participants’ oral hygiene knowledge, attitude, and practices. The same questionnaire for the pre-intervention assessment will also be used for the post-intervention assessments. However, the immediate and 6-month assessments will not include oral examinations. Figures 1 and 2 show the SPIRIT schedule and Flow diagram for the study.

**Fig 1:**
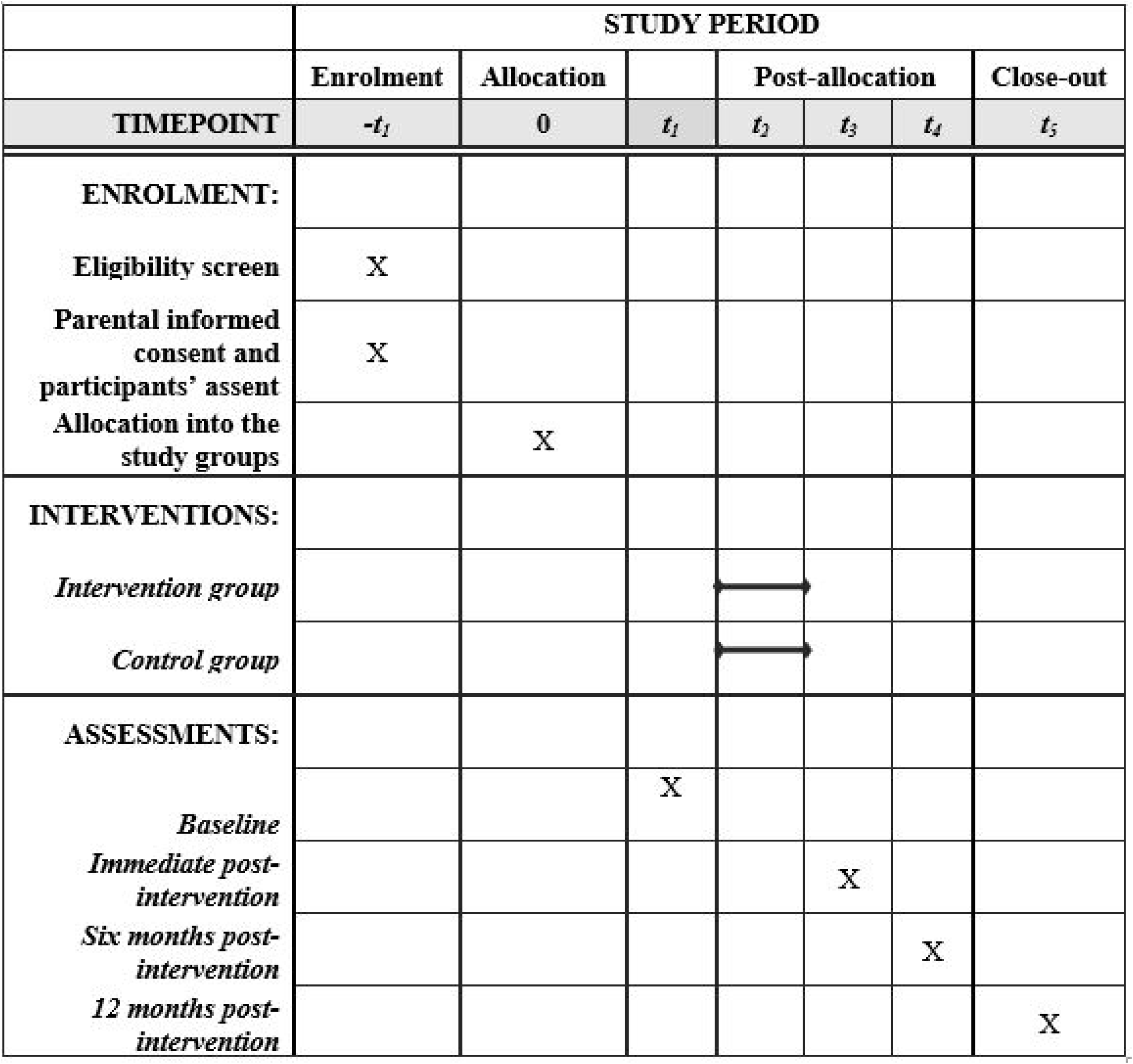
The SPIRIT schedule for the study.

**Fig 2:**
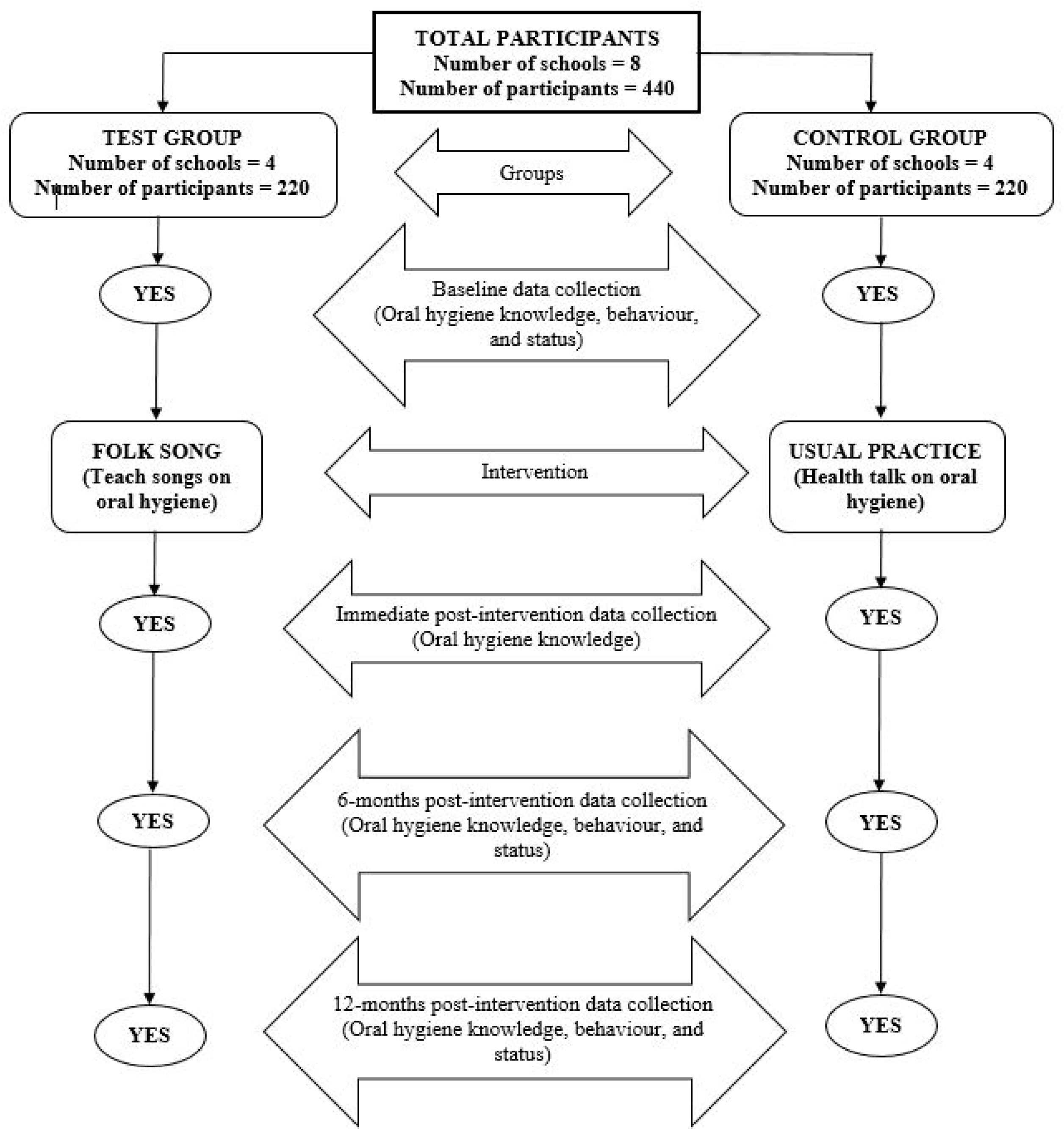
Flow diagram for the study.

## Variables

### Primary outcome variables

The participants’ oral health status will be the primary outcome variable. There would be two measures, including the caries experience, which would be based on the oral examination of the dmft/DMFT scores[29] (continuous variable) and the oral hygiene status, which would be assessed as “poor”, “fair”, and “good” (categorical variable) using the Oral Hygiene Index (Simplified) OHI-S described by Greene and Vermillion.[28]

### Secondary outcome variables

The secondary outcome variables include participants’ oral hygiene knowledge, attitude, and practices. There will be six questions assessing the participants’ knowledge. Each correct answer will be awarded a score of “1” and the wrong option “0”. Each participant’s knowledge score will be compiled over six. Similarly, there will be five questions to assess their attitude. The options will be a Likert scale (strongly disagree to strongly agree). The best response, which indicates the most positive attitude to the statement on oral hygiene, will be given the highest score of “5” while the most negative response will be scored “1”. The total attitude score for all the participants will be computed with the maximum being “25” and the minimum being “5”. The participants’ oral hygiene practices that would be assessed include twice daily brushing, the presence of fluoride and the size of dispensed toothpaste for brushing. Others are their technique of tooth brushing and how long before they changed their toothbrush.

### Independent variables

The independent variables would include the participants’ study group, sociodemographic characteristics (age, gender, neighbourhood), and school-related characteristics (school-type, class-level).

### Validity and reliability of the questionnaire

The questionnaire (Appendix I) contains mainly structured questions, with a few carefully selected open-ended questions to ensure valid responses. The questionnaire contained questions that will be easy to administer, and the importance of giving steady answers by the respondents will be emphasised. The questionnaire was face- and content-validated by dental public health experts, and they considered the questions adequate to answer the research questions and appropriate for the intended age group. The data collectors will be trained on the mode of administering the questionnaires to avoid interviewer bias. The questionnaires will be pretested among a minimum of 20 pupils in communities outside Igboora and Idere with similar relevant characteristics to the study population. The students will be asked to give feedback on the clarity of the questions and other useful information will be used to improve the questionnaire. The pretest will be done to assess the reliability of the questions, including the knowledge and attitude scales and their responses will be analysed using the test statistics of Cronbach’s reliability test. The five examiners will also individually provide oral examinations for all the 20 pupils. The examiners’ agreement on the recorded scores for the simplified oral hygiene index and dmft/DMFT will be evaluated using the Cohen Kappa statistics.

## Data management plan

### Data handling plan

The questionnaires will be checked for errors daily, and necessary corrections will be made as soon as they are discovered. Each response on the questionnaire will then be coded and recoded on a coding sheet. The codes for each response on the questionnaire will then be entered into Statistical Package for Social Science (SPSS) Version 25, after which the coded data will be checked for errors by cross-checking 10% of the data.

### Data analysis plan

Baseline data would be compared among the two study groups based on their sociodemographic characteristics and the baseline value of the outcome variables (oral hygiene status, knowledge, attitude, and practice). Subsequently, the intervention effects on the outcome variables will be assessed over the study period (baseline, immediate, 6 months and 12 months assessments). The effectiveness would also be compared between groups (test vs control) over the 12 months (baseline, 6 months, and 12-month assessment). An intention-to-treat analysis will be employed for this study.[32] A participant will remain in the originally assigned group, notwithstanding what happens during the study, such as deviation from the study protocol or loss to follow-up. The record of the last assessment will be recorded for those lost to follow-up in the subsequent assessments. The level of statistical significance for all tests will be set at p < 0.05.

## Ethical considerations

The study has been registered with the Pan African Clinical Trial Registry (PACTR202010863892797). Ethical approval was obtained from the University of Ibadan/University College Hospital Ethical Review Committee (Registration number: NHREC/05/01/2008a). The permission to carry out the study was also received from the Ministry of Education, Oyo State. The following ethical issues will be considered:

### Written informed consent

A letter to the parents or legal guardians will request their consent for their children or ward to participate in the study. An information sheet stating the details of the study will be given to them to read, after which they will be asked if they understood what the study is about and what is expected of their children/wards. Parents/legal guardians who agree to allow their children/wards to participate in the study will be asked to sign a written Informed Consent form, and their children will subsequently sign the assent form. (Appendix 2)

### Confidentiality of data

All information collected from respondents during this research will be kept confidential. All identifiable details of the participants will be separated from the coded details and collected data. The identifiable details and data entered on the computer will be password-protected and only accessible to the researchers and data entry clerks.

### Right to withdraw from the study

Study participants who wish to withdraw from the study will be allowed to do so freely at any point in time and without any negative repercussions.

### Beneficence/Non-maleficence

The study participants will benefit from the educational intervention during or immediately after the study, and the study does not pose any harm to the participants.

### Dissemination

Findings from the study will be presented at both local and international conferences and published in peer-reviewed journals.

## Discussion

A multi-disciplinary team comprising community dentists, paediatric dentists, restorative dentists and music experts was established to develop a traditional rhyme (Folk Song) to be used as a tool for oral hygiene promotion among children in two rural communities in Nigeria. The team, together with children, teachers, and parents, co-developed a simple, memorable and age-appropriate traditional song[30] as the platform for delivering oral hygiene behavioural change to children to enhance their self-efficacy with twice-daily tooth brushing.

To the best of our knowledge, this is one of the few studies to assess the effectiveness of traditional folk songs in improving children’s oral hygiene knowledge, attitude, and practices. Folk songs are very popular in Nigeria, and it is not just for entertainment but also to convey historical events and heritage and teach children about important virtues.[33,34] This study aims to use folk songs to deliver oral health education to children in two rural communities – Igboora and Idere, Oyo State, Nigeria. The burden of oral health problems is high in Nigeria, like other African countries, and those in rural areas are more affected.[13] This is one of the reasons these communities were selected for the intervention. Like other rural communities in Nigeria, access to oral health services is also limited in these communities. However, the Department of Periodontology and Community Dentistry, University College Hospital/University of Ibadan, provides basic oral health care services to these communities through the Primary Oral Health Care clinics, and some of the investigators of this research are in charge of the clinics. This relationship would facilitate a quick uptake of outcomes of this research in the subsequent planning of intervention programmes from the dental clinics in responding to oral health challenges in the study sites.

Pupils in primary schools have been targeted for this intervention because the school provides an organised setting where the participants can be taught without distractions. It also gives the best opportunity for the participants to be followed up with a lesser risk of attrition. The intervention can also be easily sustained by incorporating it into their activities during the daily morning assembly.

A school/cluster-level randomisation will be employed to avoid having participants in the intervention and control groups in the same school, which may increase the risk of spillover.[35] There is no way some participants in a school would be taught these songs, and others in the control group would not be aware if they were in the same school. In the same vein, the distance of schools will be considered when selecting the schools for the study and schools close to each other would not be allowed to be in different study groups.

This study will consider several outcomes, including knowledge, attitude, practice, and oral health status (caries and oral hygiene). This would generate empirical information on the effectiveness of folk songs on each of these outcomes separately. A possible limitation in the outcome measurement is the dmft/DMFT.

The dmft/DMFT is a non-reversible index;[29] thus, the score at baseline would not reduce at the study completion even if the participant has treated carious teeth. To address this, the condition of the teeth (decayed, missing, and filled) would be noted at baseline. Hence, if a participant visited the dental clinic to fill a decayed tooth, such would be noted as an improvement in the caries experience during the final assessment 12 months post-intervention.

## Data Availability

No datasets were generated or analysed during the current study. All relevant data from this study will be made available upon study completion.

## Trial status

The trial is yet to start the recruitment of study participants.

## Acknowledgements

The authors wish to thank FDI World Dental Federation for the funding.

## Supporting information

**S1 File: Questionnaire**

**S2 File: Parental consent and participant’s assent forms**

